# Centralized molecular testing of community-collected tongue swabs for tuberculosis screening among household contacts: a cross-sectional diagnostic performance, yield, and implementation study

**DOI:** 10.64898/2026.07.30.26359194

**Authors:** Sharon L. Olifant, Andrew Medina-Marino, Maria Pieruccini, Kuhle Fiphaza, Charl Bezuidenhout, Morten Ruhwald, Adam Penn-Nicholson, Remco P.H. Peters, P. Bernard Fourie

## Abstract

**Background:** Community-based tuberculosis screening can identify asymptomatic tuberculosis, but microbiological confirmation has been sputum-dependent. We evaluated the diagnostic performance, yield, and operational characteristics of centralized qPCR testing of community-collected tongue swabs (TS).

**Methods:** We conducted a cross-sectional study among household contacts (HHCs; ≥18 years) of individuals receiving treatment for pulmonary TB in South Africa. During household visits, health workers collected TS specimens and, where possible, sputum, which was tested in participants’ homes using Xpert MTB/RIF Ultra. TSs were transported non-refrigerated in molecular transport medium to a distant central laboratory for manual qPCR assay. The primary outcome was diagnostic performance of TS qPCR compared with sputum Xpert Ultra using paired results. Secondary outcomes were diagnostic yield by TS and operational indicators of the centralized testing workflow.

**Findings:** 909 HHCs were enrolled; median age 39 years (IQR 28–55), 532 (58·5%) were asymptomatic, 617 (67·9%) were sputum scarce. TS were collected from 901/909 (99·1%) and sputum from 292/909 (32·1%) participants. Among 271 paired TS–sputum results, TS qPCR sensitivity was 60·9% (95% CI 38·5–80·3), specificity 95·2% (91·7–97·5), and overall agreement 92·3% (88·5–95·0). Yield increased from 1·8% (95% CI 1·0–2·8) with symptom-restricted sputum testing to 3·2% (2·1–4·5) with symptom-agnostic sputum testing and 5·9% (4·5–7·7) when TS testing was implemented among sputum-scarce individuals, identifying 25 additional HHCs with positive molecular test results (86·2% increase). Operational indicators demonstrated successful implementation, with 2·3% specimen loss and valid molecular results for 93·8% who provided a TS.

**Interpretation:** Despite lower per-test sensitivity than sputum testing, TS may provide a scalable strategy for expanding microbiological screening beyond sputum-dependent pathways, thereby substantially increasing tuberculosis detection among HHCs.

**Funding:** U.S. NIH; Australian Department of Foreign Affairs and Trade; UK Foreign, Commonwealth and Development Office

**RESEARCH IN CONTEXT:** *Evidence before this study:* A growing body of evidence suggests that a large proportion of tuberculosis (TB) exists as asymptomatic disease, defined by the World Health Organization as a person with TB disease who does not report symptoms suggestive of TB during screening. However, TB detection strategies have traditionally relied on symptom-based screening of people with presumed disease who present to healthcare facilities for evaluation using sputum-based diagnostic pathways. We searched PubMed, Embase and Google Scholar for studies published in English from January 1, 2010, to July 7, 2026, using combinations of the terms “tuberculosis” or “TB”, “tongue swab”, “oral swab”, “community screening”, “household contacts”, “active case finding”, “sputum scarce”, “asymptomatic”, “subclinical”, “PCR”, “qPCR”, and “Xpert”. Community-based screening studies, including household contact investigations, have reported microbiologically detectable TB outside routine facility-based pathways, including among people without symptoms and among those unable to produce sputum. Previous studies and systematic reviews showed that tongue swab testing can detect *Mycobacterium tuberculosis* with high specificity but only moderate sensitivity, generally lower than sputum-based molecular testing and often weaker in paucibacillary and asymptomatic disease. However, the recent evaluation of the near-point-of-care Pluslife MiniDock MTB platform using sputum and tongue swabs met WHO diagnostic accuracy targets for tuberculosis detection. Recent multi-country and in-home prospective tongue swab studies suggested feasibility and diagnostic potential. However, evidence remained limited on centralized workflows, non-refrigerated transport of community-collected swabs, operational implementation indicators, and the incremental contribution of tongue swabs to microbiological case detection in sputum-scarce populations outside traditional clinic-based settings.

*Added value of this study:* This study evaluates centralized qPCR testing of community-collected tongue swabs among adult household contacts of people receiving treatment for pulmonary TB in Eastern Cape, South Africa. Relative to paired sputum Xpert MTB/RIF Ultra, tongue-swab qPCR had sensitivity of 60.9%, specificity of 95·2%, and overall agreement of 92·3%. Across screening strategies, 64 participants were microbiologically detected with TB; 51 were identified by tongue swabs and 29 by sputum, with overlap between modalities. Incorporating tongue swab testing for sputum-scarce participants identified 25 additional people with TB beyond symptom-agnostic sputum testing alone, increasing TB detection by 86·2%. Operationally, tongue swabs were collected from 99·1% of participants, the median shipment-to-receipt time was 47·5 hours, specimen loss during transport was 2·3%, and valid molecular results were generated for 93·8% of participants that provided a tongue swab specimen. To our knowledge, no previous study has combined community-based tongue swab collection, ambient transport, centralized batch qPCR testing, diagnostic performance evaluation, diagnostic-yield comparisons, and operational implementation measures in this population.

*Implications of all the available evidence:* The available evidence suggests that tongue swabs should be considered not only as an alternative specimen, but also as a programmatic access strategy for microbiological TB testing outside traditional health facilities. Centralized high-throughput tongue swab testing workflows could strengthen community-based screening by extending microbiological testing to asymptomatic and sputum-scarce populations who are not found in traditional health system settings and are often missed by sputum-dependent pathways. Priorities for future research include improving assay sensitivity, evaluating cost-effectiveness and health-system integration, assessing prospective effects on treatment initiation and transmission, and determining generalizability across populations, platforms, and settings.

## BACKGROUND

Tuberculosis (TB) control strategies have traditionally relied on symptom-based identification of people with presumed disease who present to healthcare facilities for evaluation. However, a growing body of evidence suggests that a large proportion of tuberculosis exists as asymptomatic disease, defined as person with TB disease who does not report symptoms suggestive of TB during screening.^1,2^ Population-based prevalence surveys have consistently identified asymptomatic bacteriologically confirmed TB in 36–80% of cases across settings.^3,4^ These findings are particularly relevant in South Africa, where 57·8% of bacteriologically confirmed TB cases identified through community screening would not have been detected through symptom screening alone.^5^ Asymptomatic TB is increasingly recognized as an important contributor to ongoing transmission.^6–8^ Natural history studies suggest that many individuals remain bacteriologically positive for prolonged periods without progressing to classical symptomatic disease.^9,10^ Consequently, large numbers of individuals with potentially transmissible TB are missed when relying on conventional clinic-based diagnostic pathways alone. Together, these findings highlight the need for screening approaches capable of delivering microbiological testing earlier in disease progression and beyond traditional clinic-based pathways.

Although community-based screening and active case-finding (ACF) strategies have been shown to improve identification of individuals with asymptomatic TB, microbiological confirmation remains heavily dependent on sputum-based testing.^5,11^ Recent evidence suggests that sputum scarcity is common during TB evaluation and represents a major barrier to microbiological confirmation, particularly among populations with limited ability to produce sputum.^12^ Sputum scarcity appears most common among individuals with fewer symptoms and milder disease, suggesting that sputum-based diagnostics may disproportionately limit microbiological confirmation among populations increasingly targeted through screening and case-finding strategies.^13^ In response to these limitations, the World Health Organization (WHO) recently recommended the collection and testing of tongue swab (TS) specimens for individuals that cannot produce sputum given that TS are non-invasive, acceptable, and capable of facilitating near-universal specimen collection irrespective of symptom status or sputum production.^14–17^ Multiple studies have demonstrated that molecular testing of TS specimens can identify TB across a range of clinic-based, community-based, and ACF settings.^18–20^ However, while TS may substantially expand access to microbiological TB testing, most studies have focused on diagnostic accuracy, leaving important questions regarding operational scalability and implementation largely unanswered.^21–23^

ACF and symptom-agnostic screening have re-emerged as important strategies for identifying asymptomatic TB, but their impact depends on achieving high coverage and integrating screening within feasible health-system workflows.^24–26^ Scaling these approaches requires testing architectures capable of collecting, transporting, and processing specimens efficiently from large numbers of individuals reached outside routine clinic-based care. However, existing TS studies have provided limited insight into how TS-based screening might be operationalised at population scale.^21–23^ Although near-point-of-care testing may be advantageous in some settings, large-scale screening initiatives may require testing models optimized for throughput and operational scalability. The operational characteristics of TS specimens—including non-refrigerated transport, compatibility with centralized laboratory processing, and suitability for batch-based workflows—may position them as attractive candidates for scalable community-based screening. However, whether community-collected TS specimens can be successfully integrated into centralized testing systems remains largely unknown. A critical remaining gap is whether TS can function as a specimen logistics solution rather than simply a diagnostic technology.

To address this gap, we evaluated a centralized molecular testing workflow using community-collected TS specimens among adult household contacts of individuals with pulmonary TB. We recently reported that integration of TS and near-point-of-care molecular testing can improve diagnostic access within household contact investigation (HCI). Here, we evaluated a complementary approach focused on diagnostic scalability through centralized laboratory processing of community-collected TS specimens. Specifically, we aimed to estimate the diagnostic performance of a centralized qPCR platform, quantify the contribution of TS testing to TB detection under alternative screening strategies, and evaluate operational indicators relevant to coverage, scalability, and reach to sputum-scarce populations.

## METHODS

### Study design and setting

Between June 15, 2021, and October 29, 2024, we conducted a cross-sectional diagnostic performance study embedded within a previously described prospective cohort study among household contacts (HHCs) of individuals receiving treatment for drug-sensitive pulmonary tuberculosis (DS-pTB) at 26 government health clinics in Buffalo City Metro Health District, Eastern Cape Province, South Africa.^20^ The study was designed as an exploratory early-phase evaluation to generate preliminary estimates of diagnostic performance and operational implementation characteristics under real-world conditions.

In 2023, Eastern Cape Province had an estimated TB case notification rate of 703 per 100,000 population and an HIV prevalence of 13·0% (95% CI 12·3–13·6%).^27,28^

### Participant Recruitment and Data Collection

Recruitment procedures have been described previously.^16,20^ Briefly, individuals receiving treatment for DS-pTB were consecutively approached and asked for permission to conduct a household visit. Index patients without pulmonary involvement, age <18 years, or without HHC were excluded.

During HCI, HHCs aged ≥18 years were invited to participate irrespective of TB-related symptoms. HHCs currently receiving TB treatment or tuberculosis preventive therapy (TPT), or who had completed TB treatment within the previous 6 months, were excluded. Consenting participants completed a standardized questionnaire administered using REDCap.^20^

### Tongue swab specimen collection

TS collection procedures have been described previously.^20^ Briefly, oral specimens were collected using flocked swabs (Puritan HydraFlock or Copan FLOQSwab) and placed into PrimeStore® Molecular Transport Medium (PS-MTM) (Longhorn Vaccines & Diagnostics, San Antonio, Texas, USA). During the study, specimen collection was modified from a single swab to two swabs collected into a single transport tube following emerging evidence supporting increased biomass recovery.^29^ TS specimens were transported at ambient temperature to a centralized laboratory in Pretoria, South Africa for molecular testing.

### Tongue swab processing and testing

DNA was extracted from TS specimens using commercially available extraction kits according to manufacturer instructions or protocol adaptations developed during the study. Detailed extraction procedures are provided in the Supplementary Methods.

Multiplex qPCR targeting the MTB *IS6110* and *IS1081* targets was performed using the PrimeMix® real-time MTB PCR assay (Longhorn Vaccine and Diagnostics) as previously described.^30^ Molecular testing was performed on 96-well plates. Each run included a positive, negative, and extraction control. Each participant specimen was tested in triplicate, allowing a maximum of 31 specimens to be processed per run. Staff performing laboratory procedures were blinded to the sputum Xpert Ultra result and to participant symptom status when scoring qPCR TS results.

### Sputum specimen collection and testing

After TS collection, all participants were asked to provide a spontaneous sputum specimen using standardized WHO-endorsed procedures. Sputum specimens were tested in participants’ homes using Xpert MTB/RIF Ultra cartridges (Cepheid; Sunnyvale, CA, USA) on GeneXpert Omni and EDGE (Cepheid) devices as previously described.^16,31^ Ultra-tested sputum served as the microbiological reference standard. Individuals with positive sputum Ultra test results were immediately referred for clinic-based TB treatment as previously described.^20^ Due to the research nature of our TS qPCR testing, TS results were not returned to participants or healthcare providers and did not inform clinical management.

### Outcomes

#### Primary Outcome

The primary outcome was the diagnostic performance and agreement of TS qPCR testing relative to sputum Xpert Ultra among HHCs who provided both a TS and sputum specimen. TS specimens were tested in triplicate; a TS result was considered positive if at least one replicate yielded a qPCR cycle threshold (Ct) value ≤38. Consistent with prior analyses, sputum Xpert Ultra trace results were classified as negative in the primary analysis (Table 2).

#### Secondary Outcomes

Secondary outcomes included: (1) diagnostic yield (Table 3), defined as the proportion of HHCs with any positive test result (i.e., sputum Ultra or qPCR tongue swab test) among all HHCs eligible for testing, irrespective of specimen collection or test result availability; and (2) operational implementation indicators of the centralized molecular testing workflow were organized into domains derived from key considerations outlined in the WHO Target Product Profile (TPP) for tuberculosis screening tests (Table 4).^32^ Detailed definitions of operational implementation indicators and domains are provided in the Supplementary Methods.

### Statistical Analysis

Sample size was determined pragmatically based on anticipated recruitment of household contacts over the study period, rather than formal power calculations for diagnostic accuracy, due to the study’s exploratory early-phase nature.

#### Analysis of the Primary Outcome

Diagnostic performance and agreement of TS qPCR relative to sputum Xpert Ultra were evaluated using paired TS–sputum results. Sensitivity and specificity were calculated using standard definitions.

Agreement between TS qPCR and sputum Xpert Ultra results was assessed using Cohen’s kappa statistic, McNemar’s test, and proportions of positive, negative, and overall agreement. Exact 95% CI were estimated for all diagnostic performance and agreement measures, where appropriate.

#### Analysis of Secondary Outcomes

Diagnostic yield was calculated for predefined screening and testing strategies using all enrolled household contacts (N=909) as the denominator. Whereas diagnostic performance analyses used paired TS and sputum test results with sputum Xpert Ultra as the microbiological reference standard, diagnostic yield analyses evaluated the number of participants with microbiologically detected TB under alternative screening and testing strategies using observed sputum Xpert Ultra and TS qPCR results. For each strategy, yield was calculated as the number of participants with a positive molecular test result divided by the total number of enrolled household contacts. Exact binomial 95% CIs were calculated for all yield estimates.

Operational implementation indicators were summarized using descriptive statistics, including counts, proportions, medians with interquartile ranges (IQR), and fold-change calculations.

Participant baseline characteristics were summarized using descriptive statistics, with categorical variables presented as frequencies and percentages and continuous variables as medians with interquartile ranges (IQR). Post-hoc exploratory analyses assessed agreement between TS qPCR positivity and sputum Xpert Ultra semi-quantitative categories (“high”, “medium”, “low”, “very low”, and “trace”).^33^

Descriptive analyses were conducted using R version 4.4.3 (R Foundation for Statistical Computing, Vienna, Austria), and diagnostic accuracy estimates were calculated using MedCalc Statistical Software version 23.5.2 (MedCalc Software Ltd, Ostend, Belgium) and Epitools Epidemiological Calculators (Ausvet, Fremantle, Western Australia, Australia). A *p*-value<0.05 was considered statistically significant. The 2015 Standards for the Reporting of Diagnostic accuracy studies (STARD) guidelines were followed for reporting these data (Supplemental Materials).

### Ethics and Approvals

Ethics approval was provided by the University of Pretoria (Ref. #57/2021) and University of Cape Town (Ref. #391/2021) research ethics committees. Permission was provided by the Eastern Cape Department of Health Provincial Research Committee (EC_202106_002). All participants provided written informed consent.

## RESULTS

### Study Population

Among 1,336 HHCs screened for study eligibility, 958 (71·7%) were eligible and 909 (94·9%) provided informed consent (Figure 1). The median age of enrolled HHCs was 39 years (IQR 28–55), 570 (62·7%) were female, 132 (14·5%) self-reported living with HIV, and 207 (22·8%) reported a previous history of TB (Table 1). Most participants (532/909, 58·5%) were asymptomatic. Additionally, sputum scarcity was common, affecting 399/532 (75·0%) asymptomatic and 218/377 (57·8%) symptomatic HHCs. TS and sputum specimens were collected from 901/909 (99·1%) and 292/909 (32·1%) participants, respectively, including all sputum-scarce HHCs. All participants (909/909; 100%) provided at least one specimen for microbiological testing.

**Figure 1:**
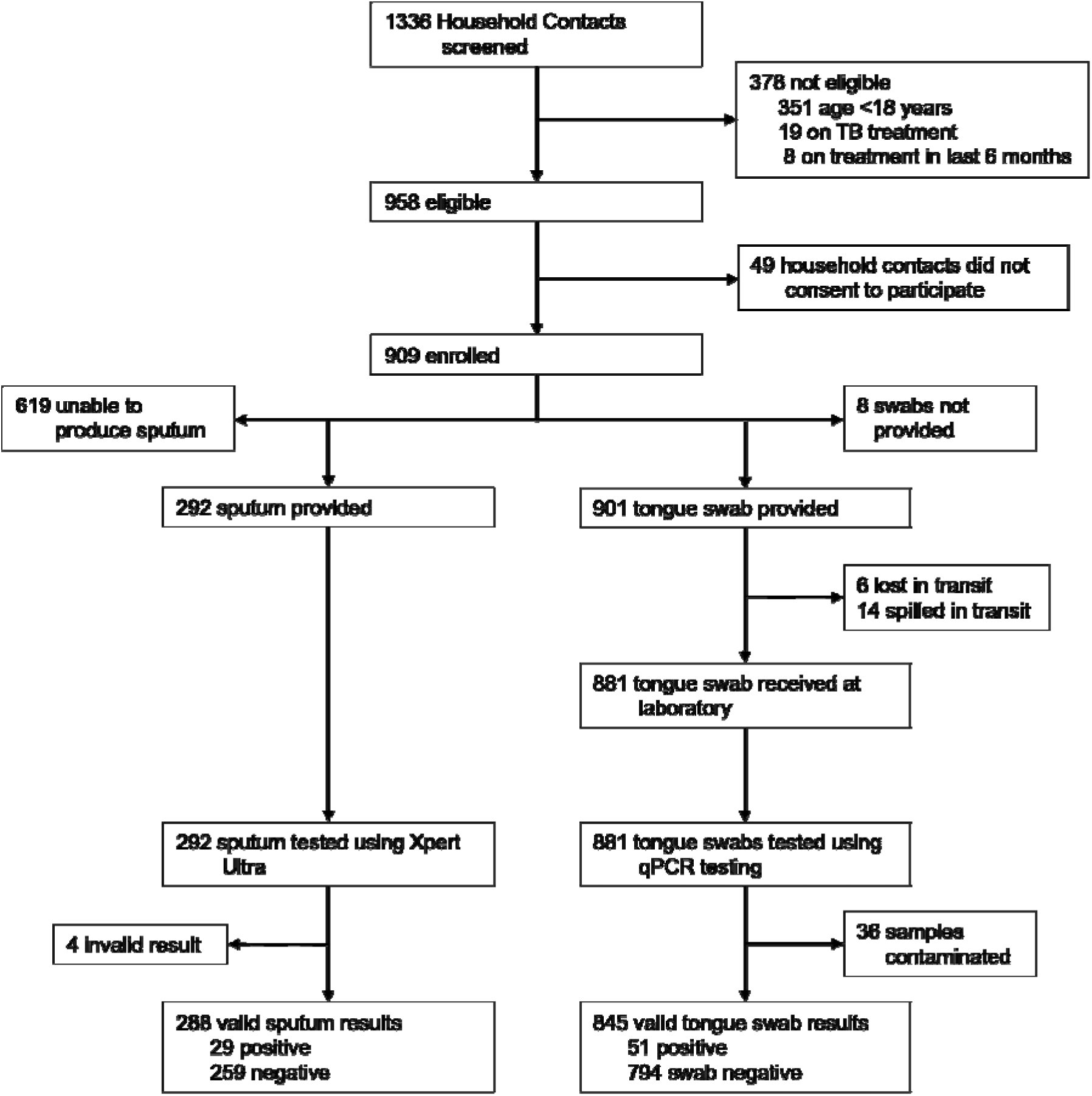
Study flow chart showing specimens collected from household contacts.

**Table 1:** Characteristics of household contacts by tuberculosis symptom status and sputum productivity.

| Characteristics | Total<br>(N=909) | Asymptomatic<br>household contact (n=532) |  | Symptomatic<br>household contact (n=377) |  |
| --- | --- | --- | --- | --- | --- |
|  |  | Sputum<br>Productive<br>(n=133) | Sputum<br>Scarce<br>(n=399) | Sputum<br>Productive<br>(n=159) | Sputum<br>Scarce<br>(n=218) |
| **Age, median [IQR]** | 39 (28-55) | 39 (29-55) | 38 (27-54) | 38 (31-54) | 41.5 (29-57) |
| **Sex** |  |  |  |  |  |
| Female | 570 (62.7%) | 71 (53.4%) | 262 (65.7%) | 90 (56.6%) | 147 (67.4%) |
| Male | 338 (37.2%) | 62 (46.6%) | 137 (34.3%) | 68 (42.8%) | 71 (32.6%) |
| Missing | 1 (0.1%) | 0 (0%) | 0 (0%) | 1 (0.6%) | 0 (0%) |
| **Race/Ethnicity** |  |  |  |  |  |
| Black | 650 (71.5%) | 81 (60.9%) | 304 (76.2%) | 102 (64.2%) | 163 (74.8%) |
| Coloured | 250 (27.5%) | 51 (38.3%) | 90 (22.6%) | 55 (34.6%) | 54 (24.8%) |
| White | 2 (0.2%) | 1 (0.8%) | 0 (0%) | 0 (0%) | 1 (0.5%) |
| Indian | 4 (0.4%) | 0 (0%) | 4 (1.0%) | 0 (0%) | 0 (0%) |
| Other | 2 (0.2%) | 0 (0%) | 0 (0%) | 2 (1.3%) | 0 (0%) |
| Missing | 1 (0.1%) | 0 (0%) | 1 (0.3%) | 0 (0%) | 0 (0%) |
| **Employment Status** |  |  |  |  |  |
| Employed | 213 (23.4%) | 29 (21.8%) | 100 (25.1%) | 40 (25.1%) | 44 (20.2%) |
| Unemployed | 475 (52.3%) | 65 (48.9%) | 208 (52.1%) | 85 (53.5%) | 117 (53.7%) |
| Retired | 65 (7.2%) | 9 (6.8%) | 31 (7.8%) | 7 (4.4%) | 18 (8.3%) |
| Student | 40 (4.4%) | 9 (6.8%) | 22 (5.5%) | 2 (1.3%) | 7 (3.2%) |
| Sick/Disabled | 16 (1.8%) | 3 (2.3%) | 5 (1.3%) | 3 (1.9%) | 5 (2.3%) |
| Other | 99 (10.9%) | 18 (13.5) | 32 (8.0%) | 22 (13.8%) | 27 (12.4%) |
| Missing | 1 (0.1%) | 0 (0%) | 1 (0.3%) | 0 (0%) | 0 (0%) |
| **History of Smoking** |  |  |  |  |  |
| Never | 476 (52.4%) | 50 (37.6%) | 261 (65.4%) | 47 (29.6%) | 118 (54.1%) |
| Current | 360 (39.6%) | 76 (57.1%) | 111 (27.8%) | 96 (60.4%) | 77 (35.3%) |
| Former | 72 (7.9%) | 7 (5.3%) | 26 (6.5%) | 16 (10.1%) | 23 (10.6%) |
| Missing | 1 (0.1%) | 0 (0%) | 1 (0.3%) | 0 (0%) | 0 (0%) |
| **Alcohol Use** |  |  |  |  |  |
| Never | 439 (48.3%) | 64 (48.1%) | 203 (50.9%) | 65 (40.9%) | 107 (49.1%) |
| Rarely (<1 day/week) | 156 (17.2%) | 19 (14.3%) | 72 (18.0%) | 24 (15.1%) | 41 (18.8%) |
| Occasionally (1-3 days/week) | 267 (29.4%) | 46 (34.6%) | 102 (25.6%) | 58 (36.5%) | 61 (28.0%) |
| Frequently (4-6 days/week) | 36 (4.0%) | 4 (3.0%) | 17 (4.3%) | 9 (5.7%) | 6 (2.8%) |
| Daily (7 days/week) | 9 (1.0%) | 0 (0%) | 3 (0.8%) | 3 (1.9%) | 3 (1.4%) |
| Missing | 2 (0.2%) | 0 (0%) | 2 (0.5%) | 0 (0%) | 0 (0%) |
| **HIV Status** |  |  |  |  |  |
| Positive | 132 (14.5%) | 15 (11.3%) | 56 (14.0%) | 25 (15.7%) | 36 (16.5%) |
| Negative | 692 (76.1%) | 100 (75.2%) | 309 (77.4%) | 119 (74.8%) | 164 (75.2%) |
| Unknown/Did not disclose | 84 (9.2%) | 18 (13.5%) | 33 (8.3%) | 15 (9.4%) | 18 (8.3%) |
| Missing | 1 (0.1%) | 0 (0%) | 1 (0.3%) | 0 (0%) | 0 (0%) |
| **Prior TB** |  |  |  |  |  |
| Never | 697 (76.7%) | 101 (75.9%) | 330 (82.7%) | 97 (61.0%) | 169 (77.5%) |
| Yes, <2 years ago | 53 (5.8%) | 9 (6.8%) | 12 (3.0%) | 19 (11.9%) | 13 (6.0%) |
| Yes, ≥2 years ago | 154 (16.9%) | 22 (16.5%) | 53 (13.3%) | 43 (27.0%) | 36 (16.5%) |
| Missing | 5 (0.6%) | 1 (0.8%) | 4 (1.0%) | 0 (0%) | 0 (0%) |
| **Times had TB in the Past<sup>†</sup>** |  |  |  |  |  |
| 1 time | 171/207 (82.6%) | 22/31 (71.0%) | 57/65 (87.7%) | 51/62 (82.3%) | 41/49 (83.7%) |
| ≥2 times | 36/207 (17.4%) | 9/31 (29.0%) | 8/65 (12.3%) | 11/62 (17.7%) | 8/48 (16.7%) |
| **TB Symptoms** |  |  |  |  |  |
| Cough | 195 (21.5%) | N/A | N/A | 110 (69.2%) | 85 (39.0%) |
| Fever | 185 (20.4%) | N/A | N/A | 81 (50.9%) | 104 (47.7%) |
| Night Sweats | 153 (16.8%) | N/A | N/A | 66 (41.5%) | 87 (39.9%) |
| Unexplained Weight Loss | 136 (15.0%) | N/A | N/A | 65 (40.9%) | 71 (32.6%) |
| **Specimen Provided** |  |  |  |  |  |
| Sputum | 292 (32.1%) | 133 (100%) | N/A | 159 (100%) | N/A |
| Tongue Swab | 901 (99.1%) | 130 (97.7%) | 399 (100%) | 154 (96.9%) | 218 (100%) |
| Both Sputum and Tongue Swab | 284 (31.2%) | 130 (97.8%) | N/A | 154 (96.9%) | N/A |
| Neither Sputum nor Tongue Swab | 0 (0%) | 0 (0%) | N/A | 0 (0%) | N/A |
| **Sputum Result** |  |  |  |  |  |
| Positive | 29/292 (9.9%) | 13/133 (9.8%) | N/A | 16/159 (10.1%) | N/A |
| Negative | 259/292 (88.7%) | 118/133 (88.7%) | N/A | 141/159 (88.7%) | N/A |
| Results Not Available | 4/292 (1.4%) | 2/133 (1.5%) | N/A | 2/159 (1.3%) | N/A |
| **Swab qPCR Test Result** |  |  |  |  |  |
| Positive | 51/901 (5.7%) | 7/130 (5.4%) | 17/399 (4.3%) | 19/154 (12.3%) | 8/218 (3.7%) |
| Negative | 794/901 (88.1%) | 121/130 (93.1%) | 352/399 (88.2%) | 127/154 (82.5%) | 194/218 (89%) |
| Results Not Available | 56/901 (6.2%) | 2/130 (1.5%) | 30/399 (7.5%) | 8/154 (5.2%) | 16/218 (7.3%) |
| **Any Positive Result** |  |  |  |  |  |
| Positive | 64/909 (7.0%) | 15/133 (11.3%) | 17/399 (4.3%) | 24/159 (15.1%) | 8/218 (3.7%) |

IQR= Interquartile range; N/A= Not applicable; <sup>¶</sup> Times had TB in the Past was restricted to participants reporting prior TB (n=207)

### Diagnostic Performance and Agreement of Tongue Swab qPCR Compared with Sputum Xpert Ultra

Among 284 participants who provided both a TS and sputum specimen, 271 had valid paired TS qPCR and sputum Xpert Ultra results available for the diagnostic performance analysis (Table 2). Within this paired-specimen subset, 23 participants had a positive sputum Xpert Ultra result in the primary analysis. In the primary analysis, in which Xpert Ultra trace results were considered negative, TS qPCR had a sensitivity of 60·9% (14/23; 95% CI 38·5–80·3) and specificity of 95·2% (236/248; 91·7–97·5) relative to sputum Xpert Ultra. Positive agreement was 57·1% (95% CI 36·5– 75·5), negative agreement was 95·7% (92·5–97·9), and overall agreement was 92·3% (88·5–95·0). Cohen’s κ was 0·53 (95% CI 0·35–0·71), consistent with moderate agreement. McNemar’s test showed no evidence of directional discordance between assays (p=0·66). In the alternative analysis, in which Xpert Ultra trace results were classified as positive, diagnostic performance estimates were similar to those from the primary analysis (Table 2, Panel A).

**Table 2:** Diagnostic performance and agreement of tongue swab qPCR relative to sputum Xpert Ultra among paired specimens (n=271)

| **Panel A: Diagnostic performance and agreement statistics by Xpert Ultra Trace negative (primary outcome) and Trace positive (alternative analysis) results** |  |  |  |
| --- | --- | --- | --- |
| **Measure** | **Ultra Trace as Negative** | **Ultra Trace as Positive** |  |
| **Diagnostic Performance** | **n/N, % (95% CI)** | **n/N, % (95% CI)** |  |
| Sensitivity | 14/23, 60.9 (38.5–80.3) | 16/29, 55.2 (35.7–73.6%) |  |
| Specificity | 236/248, 95.2 (91.7–97.5) | 232/242, 95.9 (92.5–98.0%) |  |
| **Agreement** | **Estimate (95% CI)** | **Estimate (95% CI)** |  |
| Positive agreement | 57.1% (36.5–75.5) | 58.2% (40.8–74.5%) |  |
| Negative agreement | 95.7% (92.5–97.9) | 95.3% (92.0–97.5%) |  |
| Overall agreement | 92.3% (88.5–95.0) | 91.5% (87.5–94.6%) |  |
| Cohen's Kappa | 0.53 (0.35–0.71) | 0.53 (0.37–0.70) |  |
| McNemar's test | $p=0.66$ | $p=0.68$ | |
| CI= Confidence Interval |  |  |  |
| **Panel B. Agreement of qPCR-tested tongue swabs by sputum Xpert Ultra semi-quantitative category** |  |  |  |
| **Xpert Ultra semi-quantitative category** | **Xpert Ultra Sputum Positive (n)** | **qPCR TS Positive (n)** | **Agreement (%)** |
| High | 9 | 9 | 100 |
| Medium | 3 | 2 | 66.7 |
| Low | 7 | 2 | 28.6 |
| Very Low | 4 | 1 | 25 |
| Trace | 6 | 2 | 33.3 |

In exploratory analyses, whereby TS qPCR positivity was stratified by sputum Xpert Ultra semi-quantitative category (Table 2, Panel B), TS qPCR positivity was highest among participants with high (9/9; 100%) and medium (2/3; 66·7%) semi-quantitative results. Detection was lower among participants with low (2/7; 28·6%), very low (1/4; 25%), and trace (2/6; 33·3%) results, indicating that discordance was concentrated among participants with lower semi-quantitative sputum bacillary load.

### Diagnostic Yield

Across all 909 enrolled HHCs, diagnostic yield varied substantially according to the applied screening and testing strategy evaluated (Table 3). Under current practices in which only symptomatic participants are tested using sputum, 16 participants were identified with microbiologically detected TB, corresponding to a yield of 1·8% (16/909; 95% CI 1·0–2·8%). Expanding sputum testing to all participants irrespective of symptom status increased the yield to 3·2% (29/909; 2·1–4·5%). A symptom-agnostic TS testing strategy would have identified 51 participants with a positive TS qPCR result, corresponding to a yield of 5·6% (51/909; 4·2–7·3%).

**Table 3.** Estimated diagnostic yield among all eligible household contacts under alternative testing strategies.

| Testing Strategy | People with TB (n/N) | Diagnostic Yield | 95% CI |
| --- | --- | --- | --- |
| Symptom-restricted sputum testing | 16 / 909 | 1.8% | 1.0–2.8% |
| Symptom-agnostic sputum testing | 29 / 909 | 3.2% | 2.1–4.5% |
| Symptom-agnostic tongue swab testing | 51 / 909 | 5.6% | 4.2–7.3% |
| Symptom-agnostic sputum testing plus tongue swab testing among sputum-scarce individuals<sup>s</sup> | 54 / 909 | 5.9% | 4.5–7.7% |
| Symptom-agnostic combined sputum and tongue swab testing<sup>†</sup> | 64 / 909 | 7.0% | 5.5–8.9% |

<sup>s</sup> Tongue swab testing restricted to participants unable to provide a sputum specimen
<sup>†</sup> TB was considered detected if either sputum Xpert Ultra or tongue swab qPCR was positive among participants with available test results

A strategy in which sputum testing was performed when sputum was available and TS testing was conducted among sputum-scarce participants would have identified 54 participants, corresponding to a yield of 5·9% (54/909; 4·5–7·7). When all available sputum Xpert Ultra and TS qPCR results were considered, 64 participants had at least one positive molecular result, corresponding to an overall microbiological yield of 7·0% (64/909; 95% CI 5·5–8·9). Compared with symptom-restricted sputum testing, this would have represented a four-fold increase in the number of participants identified by a positive molecular test result.

### Operational Indicators

Operational implementation indicators are presented in Table 4. TS substantially increased participant access to microbiological testing, with TS specimens collected from 901/909 (99·1%) HHCs compared with 292 (32·1%) participants who were able to expectorate sputum, representing a 3·1-fold increase in access to microbiological testing. TS collection was achieved among all 617/617 (100%) sputum-scarce HHCs, including 218/218 (100%) symptomatic and 399/399 (100%) asymptomatic individuals.

**Table 4.**
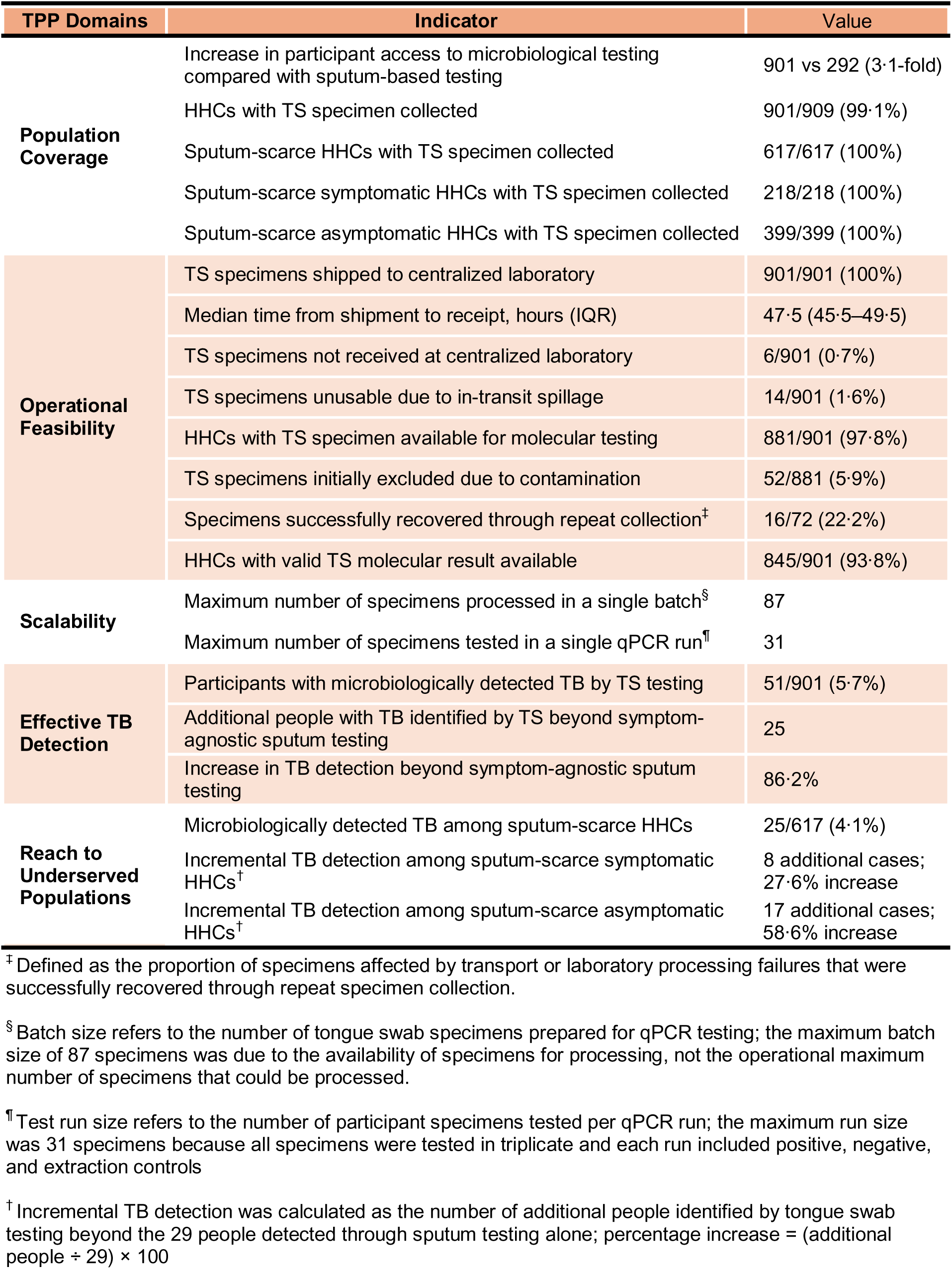
Operational implementation indicators aligned with WHO TB Screening Test Target Product Profile domains.

| TPP Domains | Indicator | Value |
| --- | --- | --- |
| **Population Coverage** | Increase in participant access to microbiological testing compared with sputum-based testing | 901 vs 292 (3.1-fold) |
|  | HHCs with TS specimen collected | 901/909 (99.1%) |
|  | Sputum-scarce HHCs with TS specimen collected | 617/617 (100%) |
|  | Sputum-scarce symptomatic HHCs with TS specimen collected | 218/218 (100%) |
|  | Sputum-scarce asymptomatic HHCs with TS specimen collected | 399/399 (100%) |
| **Operational Feasibility** | TS specimens shipped to centralized laboratory | 901/901 (100%) |
|  | Median time from shipment to receipt, hours (IQR) | 47.5 (45.5–49.5) |
|  | TS specimens not received at centralized laboratory | 6/901 (0.7%) |
|  | TS specimens unusable due to in-transit spillage | 14/901 (1.6%) |
|  | HHCs with TS specimen available for molecular testing | 881/901 (97.8%) |
|  | TS specimens initially excluded due to contamination | 52/881 (5.9%) |
|  | Specimens successfully recovered through repeat collection<sup>†</sup> | 16/72 (22.2%) |
|  | HHCs with valid TS molecular result available | 845/901 (93.8%) |
| **Scalability** | Maximum number of specimens processed in a single batch<sup>§</sup> | 87 |
|  | Maximum number of specimens tested in a single qPCR run<sup>¶</sup> | 31 |
| **Effective TB Detection** | Participants with microbiologically detected TB by TS testing | 51/901 (5.7%) |
|  | Additional people with TB identified by TS beyond symptom-agnostic sputum testing | 25 |
|  | Increase in TB detection beyond symptom-agnostic sputum testing | 86.2% |
| **Reach to Underserved Populations** | Microbiologically detected TB among sputum-scarce HHCs | 25/617 (4.1%) |
|  | Incremental TB detection among sputum-scarce symptomatic HHCs<sup>†</sup> | 8 additional cases;<br>27.6% increase |
|  | Incremental TB detection among sputum-scarce asymptomatic HHCs<sup>†</sup> | 17 additional cases;<br>58.6% increase |

<sup>†</sup> Defined as the proportion of specimens affected by transport or laboratory processing failures that were successfully recovered through repeat specimen collection.
<sup>§</sup> Batch size refers to the number of tongue swab specimens prepared for qPCR testing; the maximum batch size of 87 specimens was due to the availability of specimens for processing, not the operational maximum number of specimens that could be processed.
<sup>¶</sup> Test run size refers to the number of participant specimens tested per qPCR run; the maximum run size was 31 specimens because all specimens were tested in triplicate and each run included positive, negative, and extraction controls
<sup>†</sup> Incremental TB detection was calculated as the number of additional people identified by tongue swab testing beyond the 29 people detected through sputum testing alone; percentage increase = (additional people ÷ 29) × 100

Of 901 collected TS specimens, six (0·7%) were not received by the centralized laboratory and 14 (1·6%) were rendered unusable due to a single in-transit spillage. Median time from specimen shipment to receipt was 47·5 hours (IQR 45·5–49·5). Overall, 52/881 (5·9%) available specimens were excluded due to failed internal quality assurance steps. Of 72 specimens affected by transport or processing failures, 16 (22·2%) individuals were successfully re-engaged for repeat specimen collection. Ultimately, 845/901 (93·8%) participants from whom a TS specimen was collected had a valid molecular test result available.

TS testing microbiologically detected TB among 51/901 (5·7%) HHCs. Among sputum-scarce HHCs, TS testing microbiologically detected TB among 25/617 (4·1%) individuals, including 8/218 (3·7%) symptomatic and 17/399 (4·3%) asymptomatic participants. Overall, TS testing identified 25 additional individuals with TB beyond symptom-agnostic sputum testing, corresponding to an 86·2% increase in detection. Seventeen (68·0%) of these additional positive tests occurred among asymptomatic sputum-scarce HHCs.

### Synthesis of Study Findings

Findings from this study were synthesized with existing evidence on the spectrum of TB disease and screening pathways to develop a framework illustrating how alternative screening and testing architectures create access points for microbiological testing outside traditional health facilities and expand opportunities to detect TB across the TB disease spectrum (Figure 2).

**Figure 2.**
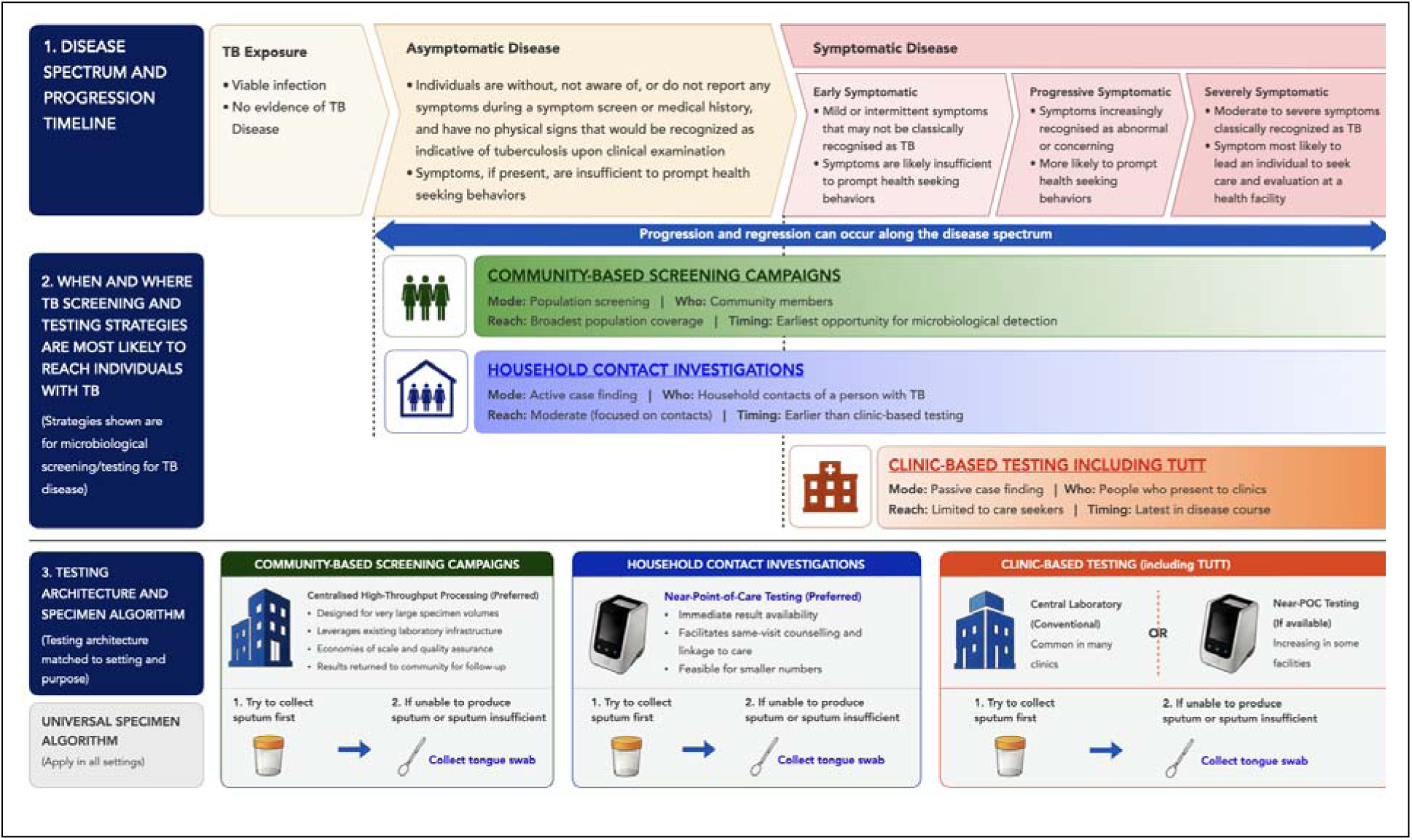
Results-informed synthesis framework illustrating how tongue-swab testing may support different TB screening and testing architectures. Framework illustrating how tongue swabs may function as a specimen-access innovation that expands microbiological testing beyond sputum-dependent pathways and enables diagnostic testing architectures to be tailored to different TB screening strategies. The figure illustrates how community-based screening campaigns, household contact investigation, and clinic-based testing are likely to reach individuals at different points along the TB disease spectrum and how distinct testing architectures may be matched to each screening context. **Upper Panel:** The upper panel depicts a simplified TB disease spectrum informed by the WHO consultation on asymptomatic TB and the ICE-TB framework.^1,2^ The framework illustrates a continuum from TB exposure through asymptomatic and symptomatic disease states. Individuals with TB exposure are generally not detectable using current microbiological tests. By contrast, asymptomatic TB may be microbiologically detectable despite the absence of recognized symptoms, while the probability of microbiological detection and healthcare-seeking generally increases across the symptomatic disease spectrum. Progression and regression may occur across disease states. **Middle Panel:** The middle panel illustrates when and where different TB screening strategies are most likely to identify individuals with TB. Community-based screening campaigns and household contact investigation may identify individuals with asymptomatic or early symptomatic disease before healthcare-seeking occurs, whereas clinic-based testing primarily identifies individuals who present for care and therefore tends to detect TB later along the disease spectrum. **Lower Panel:** The lower panel proposes a context-specific diagnostic testing architecture in which specimen collection and testing strategies are aligned with the target population, screening context, and programmatic objectives. Across all screening contexts, sputum is collected whenever possible and tongue swabs are collected from individuals unable to produce adequate sputum. However, optimal testing architectures may differ by setting. Household contact investigation may benefit from near-point-of-care molecular testing to facilitate same-visit results and linkage to care, whereas large-scale community screening initiatives may benefit from centralized high-throughput processing that leverages existing laboratory infrastructure and supports testing at scale. Clinic-based testing may utilize either centralized laboratory or near-point-of-care platforms depending on local infrastructure and programmatic needs.

## DISCUSSION

In this study evaluating centralized molecular testing of community-collected TS specimens among HHCs of individuals with pulmonary TB, we report three key findings. First, incorporation of TS testing substantially increased microbiologically detected TB and programme-level diagnostic yield among enrolled HHCs despite only moderate sensitivity of our qPCR platform relative to sputum-based molecular testing. The primary value of TS testing was therefore not superior per-test diagnostic performance but its ability to expand access to microbiological testing among sputum-scarce individuals, resulting in substantially greater diagnostic yield. Second, community-collected TSs successfully progressed through a centralized laboratory workflow characterized by near-universal specimen collection and high rates of specimen transport, laboratory receipt, and availability of valid molecular test results, demonstrating the operational feasibility of large-scale community-based TB screening approaches using TS specimens. Third, these findings highlight the importance of evaluating TB screening technologies not only according to conventional measures of diagnostic accuracy but also according to their ability to increase diagnostic coverage and effective detection across the diagnostic cascade. Together, these findings suggest that centralized processing of community-collected TS specimens may provide a scalable strategy for expanding microbiological TB screening beyond sputum-dependent testing pathways.

Evidence suggests that symptom-dependent screening and sputum-dependent testing represent sequential barriers to microbiological TB detection.^6,11,13^ In our cohort, diagnostic yield increased from 1·8% under current symptom-restricted sputum testing practices to 3·2% when sputum testing was performed irrespective of symptoms and to 5·9% when TS testing was incorporated among sputum-scarce individuals, regardless of symptom status. This symptom-agnostic screening strategy identified 25 additional individuals with TB beyond symptom-agnostic sputum testing alone, representing an 86·2% increase in people with microbiologically detectable TB. Importantly, most of these additional detection events occurred among asymptomatic sputum-scarce individuals, with TS testing identifying 17 (68·0%) of the 25 additional individuals with microbiologically detectable TB. These findings highlight the importance of distinguishing between diagnostic accuracy and diagnostic yield when evaluating TB screening technologies, as recent work has emphasized that lower-sensitivity diagnostic approaches may nonetheless achieve greater population-level case detection when they reduce losses across the diagnostic cascade and expand access to testing among individuals who would otherwise remain untested.^34,35^ Consistent with this framework, the gains observed in our study were driven primarily by expanded access to microbiological testing among sputum-scarce HHCs rather than by superior per-specimen diagnostic performance. In this context, the principal contribution of TS testing was to increase effective detection of people with microbiologically confirmed TB that could not be evaluated using sputum-based approaches.

Asymptomatic TB among sputum-scarce individuals may be systematically underestimated because burden and transmission estimates are largely derived from studies that rely on sputum-based microbiological confirmation.^7,8,36,37^ In our study, TS testing among sputum-scarce asymptomatic HHCs identified 17 additional people with microbiologically detectable TB, representing a 58·6% increase beyond symptom-agnostic sputum testing alone. These findings suggest that TS may expand access to microbiological testing among asymptomatic individuals who are unable to produce sputum, thereby revealing an under-detected reservoir of TB that remains largely invisible to symptom-dependent screening and sputum-dependent testing strategies.

A key objective of this study was to evaluate the diagnostic performance of centralized molecular TS testing. Consistent with previous evaluations of TS-based diagnostics, our assay demonstrated moderate sensitivity and high observed specificity relative to sputum-based reference testing.^32,38–40^ The observed gradient in TS sensitivity across sputum Xpert Ultra semiquantitative categories was also consistent with recent evaluations of TS-based diagnostics, including purpose-built near-point-of-care platforms, where performance declined among individuals with lower mycobacterial burden.^18^ These estimates should be interpreted in the context of the relatively small number of sputum Xpert Ultra-positive participants available for paired-specimen analysis. Consistent with recent WHO-informed modelling frameworks, the public health value of TB screening technologies depends not only on diagnostic accuracy but also on their ability to expand access to testing and reduce losses across the diagnostic cascade.^34,35^ Importantly, the WHO TPP emphasizes that screening technologies should be evaluated not only according to diagnostic accuracy but also according to their ability to achieve broad population coverage, operational feasibility, scalability, and implementation among populations not effectively reached through existing screening approaches.

Consistent with these principles, TS collection was achieved among 99·1% of HHCs, including all sputum-scarce participants, resulting in a 3·1-fold increase in access to microbiological testing compared with sputum-based testing alone. The centralized workflow demonstrated high operational feasibility, with only 0·7% of specimens not received by the centralized laboratory, 1·6% rendered unusable because of spillage, and valid molecular results available for 93·8% of participants from whom a TS specimen was collected. Successful batch processing and substantial increases in TB detection among sputum-scarce symptomatic and asymptomatic HHCs further demonstrate the scalability and public health value of this approach. Together, these findings suggest that centralized molecular TS testing may provide a practical and scalable strategy for expanding access to microbiological TB screening in high-burden settings. These findings are consistent with recent conceptual frameworks emphasizing that the public health value of TB screening technologies depends not only on diagnostic performance but also on their ability to expand access to testing and reduce losses across the diagnostic cascade.^34,35^

These findings also have implications for how microbiological TB screening programmes are designed and implemented across the TB disease spectrum (Figure 2). HCI and community-based screening campaigns reach individuals at different points along the disease spectrum than clinic-based testing programmes and therefore have distinct operational requirements. Consistent with our previous studies of in-home molecular TB testing,^20,31,41^ near-point-of-care platforms may be best suited to HCI, where testing volumes are relatively small and immediate result return, counselling, and linkage to care are important. By contrast, the successful implementation of centralized TS testing observed in this study suggests that centralized processing of community-collected specimens may be well suited to screening programmes that prioritize high testing volumes and operational scalability. In practical terms, near-POC testing may fit smaller, high-touch household-contact workflows, whereas centralized TS processing may fit larger screening programmes. Regardless of the testing architecture employed, the public health impact of microbiological screening will ultimately depend on effective return of results and linkage to care following diagnosis. Ultimately, the optimal screening and testing architecture depends on the target population, setting, availability of resources, and programmatic objectives.

To our knowledge, this is the first evaluation of centralized molecular testing of community-collected TS specimens among HHCs of individuals with pulmonary TB. It is also the first to demonstrate substantial gains in microbiologically detected TB among asymptomatic, sputum-scarce contacts through integration of TS testing within a centralized laboratory workflow. Near-universal TS collection, successful specimen transport and processing, and substantial increases in microbiologically detected TB strengthen the relevance of these findings for future community-based screening approaches. Several limitations should be considered. First, diagnostic performance estimates were derived from a relatively small number of sputa Xpert Ultra-positive participants, resulting in wide confidence intervals around sensitivity estimates. Moreover, sputum Xpert Ultra performed on a single spontaneously produced sputum specimen is an imperfect reference standard, particularly in screening populations with a high proportion of asymptomatic individuals. Because sputum reference testing was unavailable for sputum-scarce participants, the extent to which TS-positive results among these individuals represent true disease or false-positive detections could not be determined. Therefore, diagnostic yield estimates should be interpreted as measures of microbiologically detected TB under alternative screening and testing strategies rather than direct estimates of population TB burden. Second, operational implementation indicators were generated within a research-supported environment. Consequently, operational performance may differ under routine programmatic conditions. Third, the study was conducted among HHCs of individuals with TB, a population at elevated risk for disease. The diagnostic yield, operational performance, and public health impact of TS testing may differ in broader community screening settings with different epidemiologic characteristics. Despite these limitations, the study provides important evidence supporting centralized molecular TS testing as a strategy for expanding access to microbiological TB screening beyond sputum-dependent pathways.

Current efforts to expand microbiological TB screening increasingly emphasize community-based deployment of molecular diagnostics, reflecting growing recognition that substantial proportions of undiagnosed TB occur outside conventional clinic-based testing pathways. Our findings suggest that centralized processing of community-collected TS specimens may provide a complementary strategy for expanding microbiological testing at scale. Rather than relying on a single testing architecture, future screening programmes may benefit from aligning specimen collection, diagnostic platforms, and implementation strategies with the populations being reached and programmatic objectives. Future implementation studies should evaluate whether community-based TS screening programmes can translate increased access to microbiological testing into improved linkage-to-care, treatment initiation, and population-level impact under routine programmatic conditions. As efforts to identify and interrupt TB transmission increasingly focus on asymptomatic and early-stage disease, TS-based approaches may help move microbiological testing beyond sputum-dependent, clinic-based pathways and into communities that remain underserved by existing diagnostic services.

## Data Availability

Data will be made available upon request directly from the corresponding author and will include de-identified participant data (including data dictionaries) and computed variables. Additionally, code used to generate computed variable and analyse data will be provided upon request. The corresponding author and other people involved in the study will examine and review data requests. Ethical and legal implications of data sharing will be considered. Data will be shared based on the outcome of the review. The study protocol is available from the corresponding authors upon request.

## Contributors

SO contributed to study conceptualisation and laboratory methodologies, performed all laboratory-based assays, conducted formal analyses, and co-drafted the original manuscript. AM-M conceptualised the study, developed field-based methodologies, conducted formal analyses, drafted the original manuscript, supervised the study, and acquired funding. MP curated the data, conducted formal analyses, validated findings, prepared visualisations, and reviewed the manuscript. KF conducted investigations, curated data, supported project administration, and reviewed the manuscript. MR contributed resources, reviewed and edited the manuscript. CB conducted field investigations, curated data, supported project administration, reviewed and edited the manuscript. AP-N contributed to study conceptualisation, manuscript review and editing, and funding acquisition. RPHP contributed to study conceptualization, development and validation of laboratory methods, supervision and manuscript review. BF contributed to study conceptualisation, methodology, formal analysis, supervision, and funding acquisition. All authors had full access to the data, verified the integrity and accuracy of the data and analyses, agreed with the interpretation of the findings, and reviewed and approved the final manuscript. AM-M, SO and BF had final responsibility for the decision to submit for publication.

## Acknowledgments

The study was funded by the US National Institutes of Health (R01AI150485; AM-M) and FIND from the Australian Department of Foreign Affairs and Trade and the UK Foreign, Commonwealth and Development Office (AM-M and BF). The content of this study is solely the responsibility of the authors and does not necessarily represent the official views of the US National Institutes of Health, the Australian Department of Foreign Affairs and Trade, or the UK Foreign, Commonwealth and Development Office. Cepheid provided the Xpert MTB/RIF Ultra cartridges and loaned GeneXpert Omni and Edge instruments for use in the study under a collaborative agreement with FIND; no direct financial support was provided by Cepheid, and the company had no role in study design, data collection, data analysis, data interpretation, or manuscript preparation. Longhorn Vaccines and Diagnostics provided Puritan HydraFlock swabs, PrimeStore Molecular Transport Medium, and PrimeMix for qPCR assay to the University of Pretoria for research purposes. We thank the Eastern Cape Provincial Department of Health and the Buffalo City Metropolitan (BCM) Health Department, and Ms. Nondumiso Ngcelwane (BCM TB Program Manager) for their support and implementation insights toward the conducting of this study. We thank Samuel G Schumacher for conceptual support, and Grant Theron and Brooke Nichols for their critical review and feedback on drafts of this manuscript. Finally, we thank the field staff for their commitment and the study participants for their willingness to take part in this research. We acknowledge Cepheid for training, technical support, and the provision of Omni and Edge devices and Xpert MTB/RIF Ultra cartridges.

