## Supplemental Materials for "Centralized molecular testing of community-collected tongue swabs for tuberculosis screening among household contacts: a cross-sectional diagnostic performance, yield, and implementation study"

### SUPPLEMENTARY MATERIALS

#### Supplemental Methods

##### Detailed Participant Recruitment Procedures

Individuals receiving treatment for drug-sensitive pulmonary tuberculosis (DS-pTB) were identified through participating government health clinics and consecutively approached for recruitment. Index patients were excluded if they did not have pulmonary involvement, were younger than 18 years of age, lived alone, or reported no household contacts.

For eligible index patients, study staff generated a roster of household members and scheduled a household visit, typically within 3–4 days of enrollment. During household visits, study staff obtained permission from a household representative to enter the household and conduct study procedures. Refusal by the household representative terminated further household engagement. Individual participation was voluntary and written informed consent was obtained from all participants before study procedures.

Per South African national tuberculosis guidelines, household contacts younger than 18 years of age and individuals declining study participation were referred to local healthcare facilities for tuberculosis screening and management services.

##### Detailed Tongue Swab Collection Procedures

Tongue swab (TS) specimens were collected by trained study staff using flocked swabs and PrimeStore Molecular Transport Medium (PS-MTM; Longhorn Vaccines & Diagnostics, Texas, USA). Initially, a single Puritan HydraFlock® swab (Puritan Medical Products, Guilford, Maine, USA) was collected from each participant. The swab was rolled across the soft palate from left to right approximately 6–7 times (approximately 10 seconds), followed by 6–7 passes across the dorsal surface of the tongue (approximately 10 seconds). The swab shaft was then broken and the swab tip placed into a tube containing 1.5 mL PS-MTM.

During study implementation, the collection protocol was modified following reports demonstrating increased biomass recovery using Copan FLOQSwabs® compared with alternative swab types.[1] Under the revised protocol, two Copan FLOQSwabs® (Copan Diagnostics, Murrieta, California, USA) were collected sequentially and placed into a single PS-MTM tube. All subsequent laboratory processing was performed on the combined specimen.

PS-MTM was selected because it inactivates infectious organisms while preserving nucleic acids at ambient temperature, thereby facilitating transport without cold-chain requirements.

##### Specimen Storage and Transport

Following collection, TS specimens were individually packaged in leakproof transport bags and maintained at ambient temperature.

Specimens were accumulated at the field site and shipped in batches, generally once weekly and typically when approximately 20 or more specimens had been collected. Specimens were transported using commercial courier services from East London, Eastern Cape Province, South Africa, to the centralized laboratory in Pretoria, Gauteng Province, South Africa, a transport distance of approximately 1200 km.

Dates and times of specimen collection, shipment, receipt, testing, and result availability were recorded to permit evaluation of operational implementation indicators associated with specimen transport and centralized processing.

##### Detailed DNA Extraction Procedures

PrimeXtract Extraction Protocol

At study initiation, DNA extraction was performed using PrimeXtract® DNA Extraction Kits (Longhorn Vaccines & Diagnostics, Texas, USA) according to manufacturer instructions.

Briefly, aliquots of PS-MTM specimen were processed using the PrimeXtract extraction workflow and purified DNA was subsequently used for qPCR testing.

QIAamp Extraction Protocol

Following discontinuation of the PrimeXtract extraction kits by the manufacturer, DNA extraction was transitioned to QIAamp DNA Mini Kits (Qiagen, Venlo, Netherlands).

Because PS-MTM chemically lyses mycobacterial organisms during specimen collection and storage, several modifications to the standard QIAamp protocol were implemented to maximize DNA recovery:

1. The standard QIAamp lysis step was omitted.
2. The entire 1.5 mL PS-MTM specimen volume was processed.
3. To accommodate the specimen volume, two sequential 750 µL aliquots were loaded onto the same spin column and centrifuged at 6000 × g for one minute following each loading step.
4. Elution time was increased from one minute to three minutes at room temperature before centrifugation to maximize DNA recovery.

Purified DNA was stored according to manufacturer recommendations until molecular testing.

#### Detailed qPCR Testing Procedures

Molecular testing was performed using the PrimeMix® real-time MTB PCR assay (Longhorn Vaccines & Diagnostics, Texas, USA), a multiplex qPCR assay targeting the *Mycobacterium tuberculosis* insertion sequences IS6110 and IS1081.[27]

Testing was conducted on 96-well PCR plates. Each plate included:

- One positive amplification control
- One negative amplification control
- One extraction control

Participant specimens were tested in triplicate. A TS specimen was classified as positive if at least one of three replicates produced a cycle threshold (Ct) value ≤38 for either MTB target.

Because three wells were reserved for quality-control procedures and each participant specimen was tested in triplicate, a maximum of 31 participant specimens could be processed per qPCR run under the study protocol.

#### Diagnostic Yield Definitions

Diagnostic yield was calculated for predefined screening and testing strategies using all enrolled household contacts (N=909) as the denominator.

The following strategies were evaluated:

Strategy 1: Symptom-Restricted Sputum Testing

Current standard practice in many settings in which sputum testing is restricted to participants reporting TB-related symptoms.

Strategy 2: Symptom-Agnostic Sputum Testing

Sputum testing performed irrespective of symptom status among participants able to provide sputum.

Strategy 3: Symptom-Agnostic Tongue Swab Testing

TS testing performed irrespective of symptom status among participants providing a TS specimen.

Strategy 4: Combined Sputum–Tongue Swab Testing

Symptom-agnostic testing in which sputum was tested when available and TS testing was performed among individuals unable to produce sputum.

Strategy 5: Any Positive Molecular Test

Participants classified as positive if either sputum Xpert Ultra or TS qPCR yielded a positive result.

#### Operational Implementation Indicator Definitions

Operational implementation indicators were organized into domains informed by key considerations described in the WHO Target Product Profile (TPP) for tuberculosis screening tests.[2]

Population Coverage

Indicators describing access to microbiological testing within the study population, including specimen collection coverage and access to any microbiological testing modality.

Operational Feasibility

Indicators describing successful completion of workflow steps required for centralized testing, including specimen transport, specimen receipt, specimen integrity, test completion, recovery of affected specimens, and generation of valid molecular results.

Scalability

Indicators describing the capacity of the workflow to support centralized high-throughput testing, including batch processing characteristics and laboratory throughput.

Effective Case Detection

Indicators describing microbiologically detected tuberculosis cases identified through TS testing and the incremental contribution of TS testing to overall case detection.

Reach to Underserved Populations

Indicators describing the ability of TS testing to identify microbiologically detected TB among populations not effectively reached through sputum-dependent testing, including sputum-scarce participants and asymptomatic household contacts.

#### Supplementary Statistical Methods

Alternative Trace Classification Analysis

The primary analysis classified Xpert Ultra Trace results as negative, consistent with prior analyses and the study's primary analytic plan.

A prespecified alternative analysis classified Trace results as positive. Diagnostic performance and agreement statistics were recalculated under this alternative classification to assess robustness of findings to different Trace result interpretations.

Exploratory Semi-Quantitative Analysis

Post-hoc exploratory analyses evaluated agreement between TS qPCR positivity and sputum Xpert Ultra semi-quantitative categories (high, medium, low, very low, and trace).[3] These analyses were descriptive and intended to explore potential relationships between mycobacterial burden and TS detection.

#### Standards for Reporting Diagnostic Accuracy Studies (STARD) 2015 Checklist

| Section & Topic | No | Item | Reported on page # |
|---|---|---|---|
| **TITLE OR ABSTRACT** | | | |
| | **1** | Identification as a study of diagnostic accuracy using at least one measure of accuracy (such as sensitivity, specificity, predictive values, or AUC) | 1 and 2 |
| **ABSTRACT** | | | |
| | **2** | Structured summary of study design, methods, results, and conclusions (for specific guidance, see STARD for Abstracts) | 2 |
| **INTRODUCTION** | | | |
| | **3** | Scientific and clinical background, including the intended use and clinical role of the index test | 5-6 |
| | **4** | Study objectives and hypotheses | 6 |
| **METHODS** | | | |
| *Study design* | **5** | Whether data collection was planned before the index test and reference standard were performed (prospective study) or after (retrospective study) | 7 |
| *Participants* | **6** | Eligibility criteria | 7 |
| | **7** | On what basis potentially eligible participants were identified (such as symptoms, results from previous tests, inclusion in registry) | 7 |
| | **8** | Where and when potentially eligible participants were identified (setting, location and dates) | 7 |
| | **9** | Whether participants formed a consecutive, random or convenience series | 7 |
| *Test methods* | **10a** | Index test, in sufficient detail to allow replication | 8 |
| | **10b** | Reference standard, in sufficient detail to allow replication | 8 |
| | **11** | Rationale for choosing the reference standard (if alternatives exist) | 8 |
| | **12a** | Definition of and rationale for test positivity cut-offs or result categories of the index test, distinguishing pre-specified from exploratory | 8 |
| | **12b** | Definition of and rationale for test positivity cut-offs or result categories of the reference standard, distinguishing pre-specified from exploratory | 8 |
| | **13a** | Whether clinical information and reference standard results were available to the performers/readers of the index test | 8 |
| | **13b** | Whether clinical information and index test results were available to the assessors of the reference standard | 8 |
| *Analysis* | **14** | Methods for estimating or comparing measures of diagnostic accuracy | 8-9 |
| | **15** | How indeterminate index test or reference standard results were handled | 8-9 |
| | **16** | How missing data on the index test and reference standard were handled | 8-9 |
| | **17** | Any analyses of variability in diagnostic accuracy, distinguishing pre-specified from exploratory | 8-9 |
| | **18** | Intended sample size and how it was determined | 8 |
| **RESULTS** | | | |
| *Participants* | **19** | Flow of participants, using a diagram | 18 |
| | **20** | Baseline demographic and clinical characteristics of participants | 11, 20-12 |
| | **21a** | Distribution of severity of disease in those with the target condition | 11, 20-21 |
| | **21b** | Distribution of alternative diagnoses in those without the target condition | N/A |
| | **22** | Time interval and any clinical interventions between index test and reference standard | 8 |
| *Test results* | **23** | Cross tabulation of the index test results (or their distribution) by the results of the reference standard | 11, 22 |
| | **24** | Estimates of diagnostic accuracy and their precision (such as 95% confidence intervals) | 11, 22 |
| | **25** | Any adverse events from performing the index test or the reference standard | N/A |
| **DISCUSSION** | | | |
| | **26** | Study limitations, including sources of potential bias, statistical uncertainty, and generalisability | 15 |
| | **27** | Implications for practice, including the intended use and clinical role of the index test | 16 |
| **OTHER INFO** | | | |
| | **28** | Registration number and name of registry | N/A |
| | **29** | Where the full study protocol can be accessed | 17 |
| | **30** | Sources of funding and other support; role of funders | 4, 9-10, 17-18 |
